# Comparison of Large Language Models’ Performance on Neurosurgical Board Examination Questions

**DOI:** 10.1101/2025.02.20.25322623

**Authors:** Nicholas S. Andrade, Surya Donty

## Abstract

**Background:** Multiple-choice board examinations are a primary objective measure of competency in medicine. Large language models (LLMs) have demonstrated rapid improvements in performance on medical board examinations in the past two years. We evaluated five leading LLMs on neurosurgical board exam questions.

**Methods:** We evaluated five LLMs (OpenAI o1, OpenEvidence, Claude 3.5 Sonnet, Gemini 2.0, and xAI Grok2) on 500 multiple-choice questions from the Self-Assessment in Neurological Surgery (SANS) American Board of Neurological Surgery (ABNS) Primary Board Examination Review. Performance was analyzed across 12 subspecialty categories and compared to established passing thresholds.

**Results:** All models exceeded the threshold for passing, with OpenAI o1 achieving the highest accuracy (87.6%), followed by OpenEvidence (84.2%), Claude 3.5 Sonnet (83.2%), Gemini 2.0 (81.0%) and xAI Grok2 (79.0%). Performance was strongest in Other General (97.4%) and Peripheral Nerve (97.1%) categories, while Neuroradiology showed the lowest accuracy (57.4%) across all models.

**Conclusions:** State of the art LLMs continue to improve, and all models demonstrated strong performance on neurosurgical board examination questions. Medical image analysis continues to be a limitation of current LLMs. The current level of LLM performance challenges the relevance of written board examinations in trainee evaluation and suggests that LLMs are ready for implementation in clinical medicine and medical education.

## Introduction

Medical board certification relies on multiple-choice questions to provide an objective, efficient way to assess knowledge at scale [3], [7]. The American Board of Neurological Surgery adopted this approach in 1962 [17] and continues to use it today, requiring candidates to pass a 100-question Neuroanatomy exam and a 375-question primary examination covering basic science and clinical knowledge.

The 2017 development of the transformer architecture fundamentally changed how machines process language [16]. This innovation allowed models to analyze relationships between words across long text sequences, enabling more sophisticated understanding and reasoning. The impact has been dramatic. Early large language models in 2020-21 produced confident but often incorrect answers [11]. Today’s models consistently demonstrate human-level performance across professional examinations in medicine, law, and PhD-level mathematics and science [4], [8], [2], [19].

Initial studies of LLM performance on neurosurgery board examinations showed promising results. In early 2023, ChatGPT achieved a 73.4% score on mock board examinations, while its successor GPT-4 scored 83.4%, significantly outperforming both ChatGPT and human test-takers [1]. A study of the European Board Examination in Neurological Surgery found that commercial LLMs could pass the written portion, though they struggled with image-based questions [15]. Most recently, a systematic review found that GPT-4 achieved passing scores on 26 of 29 medical board examinations across specialties [12].

We evaluated five leading LLMs on 500 neurosurgical board examination questions to understand their current capabilities and implications for medical education. Our results suggest both opportunities and challenges in how we train and assess medical professionals in an AI-augmented future.

## Methods

The Congress of Neurological Surgeons Self-Assessment in Neurological Surgery (SANS) Primary Board Examination Review is an educational resource designed to prepare neurosurgical residents for board certification and practitioners for continuing medical education [5]. These questions are behind a paywall, reducing the chances that they formed part of the training data for any of the LLMs studied. We used Exam 1, which comprises 500 questions in the following categories: Vascular, Pain, Spine, Peripheral Nerve, Fundamentals, Trauma, Other General, Neuroradiology, Neuropathology, Functional, Tumor, and Pediatrics.

We evaluated five leading LLMs: OpenAI o1 [8], Anthropic’s Claude 3.5 Sonnet [2], Google’s Gemini 2.0 [6], xAI’s Grok2 [18] and OpenEvidence [13], a specialized medical answer engine.

Each question’s text and answer choices were entered into the models’ standard web interfaces. For questions containing images, we uploaded these when supported by the model’s interface. The model responses were then entered into the SANS website to validate the accuracy and record the results of the sections.

Statistical analysis compared each model’s performance to the standard passing threshold using the one-sample binomial test. Comparisons between models used the Z test for two proportions (significance: p*<*0.05).

## Results

All models achieved greater accuracy than the 70% passing threshold (p*<*0.05). OpenAI o1 significantly outperformed xAI Grok2 (p = 0.0003), Gemini 2.0 (p = 0.0041) and Claude 3.5 (p = 0.0488), while OpenEvidence also significantly outperformed xAI Grok2 (p = 0.0338); all other model comparisons did not show statistically significant differences in performance.

**Table 1.** Performance of Different Models on Neurosurgical Board Examination Questions.

| Model | Accuracy |
| --- | --- |
| OpenAI o1 | 87.6% |
| Open Evidence | 84.2% |
| Claude 3.5 Sonnet | 83.2% |
| Gemini 2.0 | 81.0% |
| xAI Grok2 | 79.0% |

The models performed significantly above the mean in five sections (Other General, Peripheral Nerve, Functional, Fundamentals, and Spine) and significantly below the mean in two sections (Neuroradiology and Vascular).

**Table 2.** Average Accuracy of Models Across Sections.

| Section | Accuracy |
| --- | --- |
| Other General | 97.4% |
| Peripheral Nerve | 97.1% |
| Functional | 96.2% |
| Fundamentals | 95.6% |
| Spine | 89.3% |
| Pain | 88.0% |
| Tumor | 86.3% |
| Pediatrics | 84.6% |
| Trauma | 80.6% |
| Vascular | 77.2% |
| Neuropathology | 76.7% |
| Neuroradiology | 57.4% |

## Discussion

All tested LLMs exceeded the neurosurgical board examination passing threshold of 70%. Lower performance in Neuroradiology reflects current limitations in LLM image processing capabilities [1], though specialized computer vision models already match or exceed human performance in tasks like mammogram interpretation [10] and tumor segmentation [9]. Some advanced medical AI systems like Google’s Med-PaLM [14] remain restricted from public testing, suggesting our results may underestimate AI’s current potential in medicine.

The strong performance of LLMs challenges the relevance of multiple-choice examinations in medical education. When machines can consistently outperform humans on tests meant to assess medical knowledge, evaluation methods should be reconsidered. The traditional acceptance of 60-70% passing scores implies that physicians graduate with significant knowledge gaps. If the tested knowledge is truly essential, LLMs should be used to ensure comprehensive understanding rather than accepting these gaps. LLMs arrive at a time of controversy for MCQ assessments like the SAT, MCAT and Step 1: are they meant to confirm basic competency or identify talent for desirable careers?

LLMs could transform medical education through personalized learning paths, on-demand explanations, and simulation of rare cases. Rather than spending years memorizing information that LLMs can instantly recall, medical training should focus on skills that AI cannot yet replicate. Developing objective measures of technical expertise is an obvious goal for procedural specialties.

As clinical tools, LLMs could provide always-on analysis of medical documentation, flag potential diagnoses, suggest relevant literature, and identify gaps in clinical workup. EHR integration and clear frameworks for liability remain unresolved problems. Most importantly, these systems must enhance rather than disrupt physician cognitive processes, providing insights at appropriate moments without causing alert fatigue or cognitive overload.

## Data Availability

All data produced are available online at GitHub.com/nsa122/LLM-NS-boards

